# Impact of Retinopathy and Systemic Vascular Comorbidities on All-Cause Mortality

**DOI:** 10.1101/2021.01.19.21249177

**Authors:** Zhuoting Zhu, Xianwen Shang, Wei Wang, Jason Ha, Yifan Chen, Jingyi He, Wenyi Hu, Xiaohong Yang, Mingguang He

## Abstract

**Aims/hypothesis:** To investigate the joint effects of retinopathy and systemic vascular comorbidities on mortality.

**Methods:** This study included 5703 participants (≥40 years old) from the 2005-2008 National Health and Nutrition Examination Survey. The Early Treatment Diabetic Retinopathy Study grading scale was used to evaluate the retinopathy status. Systemic vascular comorbidities included diabetes mellitus (DM), high blood pressure (HBP), chronic kidney disease (CKD) and cardiovascular disease (CVD). Time to death was calculated as the time from baseline to either the date of death or censoring (December 31^st^, 2015), whichever came first. Risks of mortality were estimated using Cox proportional hazards models.

**Results:** After adjusting for confounders, the presence of retinopathy predicted higher all-cause mortality (hazard ratio [HR], 1.40; 95% confidence interval [CI], 1.09-1.81). The all-cause mortality among participants with both retinopathy and systemic vascular comorbidities including DM (HR, 1.63; 95% CI, 1.06-2.50), HBP (HR, 1.46; 95% CI, 1.03-2.08), CKD (HR, 1.71; 95% CI, 1.24-2.35) and CVD (HR, 1.88; 95% CI, 1.19-2.96) was significantly higher than that among those without either condition.

**Conclusions/interpretation:** In this prospective study, individuals with retinopathy had increased all-cause mortality. The joint effects of retinopathy and major systemic vascular comorbidities increased the all-cause mortality further, suggesting that more extensive vascular risk factor assessment and management are needed to detect the burden of vascular pathologies and improve long-term survival in individuals with retinopathy.

**Research in context:** *What is already known about this subject?:* Retinopathy has been recognized as an independent risk factor for mortality.

*What is the key question?:* What are the joint effects of retinopathy and systemic vascular comorbidities (including diabetes mellitus, hypertension and chronic kidney disease and cardiovascular disease) on mortality?

*What are the new findings?:* Consistent evidence on the increased risk of mortality among individuals with retinopathy was noted in a large sample of middle-aged and older adults. The co-occurrence of retinopathy and systemic vascular conditions (diabetes mellitus, hypertension and chronic kidney disease and cardiovascular disease) further increased all-cause mortality independent of other covariates.

*How might this impact on clinical practice in the foreseeable future?:* Individuals with retinopathy may benefit from a comprehensive vascular assessment. Intensive vascular risk reduction is needed in the management of patients with retinopathy and and micro- or macrovascular disorders. Highlighted the importance of retinopathy screening using retinal imaging for identifying individuals at high risk of mortality, particularly among individuals with systemic vascular comorbidities.

## Introduction

Retinopathy commonly refers to a spectrum of signs on the fundus (e.g. microaneurysms, soft/hard exudates, and/or retinal hemorrhage) that are common in the elderly, even in those without diabetes.^1^ A recent pooled analysis of 22,896 individuals with diabetes reported that nearly 35% had retinopathy.^2^ Furthermore, previous evidence suggested that up to 15% of non-diabetic patients exhibited clinical signs of retinopathy.^3-7^

Previous literature has consistently described significant associations between retinopathy and systemic vascular comorbidities, including diabetes mellitus (DM),^5^ high blood pressure (HBP),^8^ chronic kidney disease (CKD)^6, 9-11^ and cardiovascular disease (CVD).^12-15^ The presence of retinopathy has been reported to be an independent risk factor for mortality.^16-27^ A recent meta-analysis of 20 observational studies reported that individuals with retinopathy had a two-to four-fold increase in their all-cause mortality independent of other potential risk factors.^28^

To date, only a few studies have investigated the joint effects of retinopathy and systemic vascular comorbidities (such as DM,^16^ CKD,^16, 29-31^ and clinical cerebrovascular disease^16^) on mortality. These previous studies were subject to selection bias, underestimation of retinopathy, short duration of follow-up and a lack of investigation of the effects of concomitant macrovascular disorders. Given that the retina is readily viewable via non-invasive retinal imaging and that there is a well-established association between retinopathy and systemic vascular comorbidities, clarifying the precise effects of the co-occurrence of retinopathy and systemic vascular comorbidities on the risk of mortality is of great significance.

The National Health and Nutrition Examination Survey (NHANES) is a large nationally representative multiethnic sample of U.S. adults, with a standardized grading protocol for retinopathy using photographs of two eyes, availability of death records from 2005 through 2015, information on macrovascular disorders and inclusion of a comprehensive range of covariates. Therefore, the analysis based on the NHANES address previous limitations and provide more robust estimates of impacts of retinopathy and systemic vascular comorbidities on mortality.

## Methods

### Sample and Population

The NHANES dataset from 2005 to 2008 was used for the current analysis. NHANES is a nationally representative survey of the non-institutionalized U.S. civilian population that includes in-person interviews and extensive clinical examinations. As described in detail elsewhere, the NHANES uses a multistage design to select participants from strata and proportions of minority populations.^32^ Since 1999, the NHANES has released a dataset every two years. Because of the availability of retinal imaging, two NHANES cycles (2005-2006 and 2007-2008) were merged. In total, 5,703 out of 6,797 individuals aged 40 years and older were included for the current analysis. 1,093 participants were excluded due to missing information on the grading of retinopathy for both eyes. One participant was excluded because of the lack of mortality data.

The research adhered to the tenets of the Declaration of Helsinki. Written informed consent was given by every participant, and the NHANES was conducted in accordance with ethical standards. Because data used in this analysis were publicly available and de-identified, the present study received exemption from review by the institutional review board.

### Evaluation of Retinopathy

The Canon CR6-45NM Ophthalmic Digital Imaging System and Canon EOS 10D digital camera (Canon USA Inc., One Canon Park, Melville, New York) were used to capture non-mydriatic retinal photographs. Two digital images per eye (one for the macula and one for the optic nerve) were taken in an almost completely dark room. Based on a modification of the Airlie House classification system,^33, 34^ graders in the University of Wisconsin Ocular Epidemiologic Reading Center, Madison assessed the fundus photographs. Details of image capture and grading of fundus photographs have been described in an earlier study.^35^ The Early Treatment Diabetic Retinopathy Study (ETDRS) grading scale was used to evaluate the retinopathy severity.^33^ Participants were classified as having no retinopathy (level 10-13), minimal-to-mild non-proliferative retinopathy (minimal-to-mild NPR, level 14-31), moderate-to-severe retinopathy non-proliferative retinopathy (moderate-to-severe NPR, level 41-51) and proliferative retinopathy (PR, level ≥ 60) based on the eye with the worse

retinopathy level.

### Assessment of Systemic Vascular Comorbidities

Systemic vascular comorbidities, including DM, HBP, CKD and self-reported history of CVD, were assessed. Identification of DM was based on self-reported medical history, use of insulin or antihyperglycaemic medications, or a glycosylated hemoglobin level exceeding 6.5%.^36^ Identification of HBP was based on a self-reported physician diagnosis, use of antihypertensive medications, or the average of three measurements of systolic ≥140 mmHg) and/or the average of three measurements of diastolic blood pressure ≥90 mmHg). An estimated glomerular filtration rate (eGFR) of less than 60 ml/min/1.73 m^2^ was used to as the definition for CKD.^37^ A self-reported physician diagnosis of coronary heart disease, myocardial infarction, congestive heart failure, angina or stroke was used to identify the self-reported history of CVD.

### Mortality Data

Mortality status was determined by NHANES linked National Death Index (NDI) public-access files through December 31, 2015. Based on a probabilistic matching algorithm, mortality was ascertained from the NDI.^38^ This linkage was performed by matching participants’ personal identification information, such as name, sex, date of birth, social security number, between the NHANES and the NDI datasets. Participants not found to be deceased were considered to be alive. The time between the date of the interview and the date of death or end-of-study censoring (the 31^st^ of December, 2015), whichever came first, was calculated as the duration of follow-up.

### Covariates

Ethnicity was classified into four groups: non-Hispanic white, non-Hispanic black, Mexican American, and other. Two categories of marital status included unmarried or other, and married or lived with a partner. Education level was classified into two groups: did not complete a high school degree, and completed at least a high school degree. The poverty-income ratio (PIR) was categorized into two groups: < 1.00 and ≥ 1.00 (poverty threshold)

respectively. Smoking status was classified into two groups: never, and former or current smokers. Drinking status was classified into two groups: abstainer or former drinker, and current drinker. Duration of diabetes was obtained from the questionnaire. Identification of hypercholesterolaemia was based on the total cholesterol level (≥240 mg/dL) or the use of antihyperlipidemic agents. Body mass index (BMI) was a value derived from the weight in kilograms divided by the height in meters squared. A high C-reactive protein (CRP) was defined as CRP level ≥1 mg/dL. Walking disability was based on self-reported responses to the questionnaire or the requirement of special equipment to aid walking. Health status was dichotomously classified into poor or fair, and good or excellent.

### Statistical Analysis

The complex, stratified design of NHANES was taken into account for all analyses. The means and standard errors (SEs), or numbers and weighted proportions were used to present the baseline characteristics of the study population where appropriate. The design-adjusted t-test or the Rao-Scott Pearson χ^2^ were used to compare distributions for continuous or categorical variables. We used Kaplan-Meier estimates to generate plots of survival curves among participants with retinopathy and concomitant systemic vascular disorders, and log-rank tests to compare survival distributions among these groups. We used Cox proportional hazards regression models to estimate hazard ratios (HRs) and 95% confidence intervals (CIs) for survival. Furthermore, Cox proportional hazard regression models were used to model the relationship between all-cause mortality and exposures of retinopathy and systemic vascular diseases according to a reference group of participants without retinopathy and systemic vascular comorbidity. We also examined additional effects from vascular comorbidities on all-cause mortality according to a reference group of those with either retinopathy or systemic vascular comorbidity. In order to address the potential non-response bias, we have built models for the probability of inclusion and using inverse probability weighting in sensitivity analysis. We tested the proportional hazard assumption for each variable in the Cox regression models by creating the time-dependent covariate (interaction between the variable and survival time), with a p-value < 0.05 for the time-dependent covariate indicating a violation of the assumption. We found that all variables included in the Cox regression models were valid. Collinearity among variables was tested using the variance inflation factor (VIF) procedure, and the average value was 1.28 in the present analysis. We used Stata (ver. 14.0; StataCorp., College Station, TX) to perform all analyses. A p-value of less than 0.05 was considered statistically significant.

## Results

In total, there were 6,797 individuals aged 40 years and older from the NHANES 2005-2008. Of these participants, 5,703 were included in the current analysis due to missing data on fundus photography (n=969), ungradability of photographs for both eyes (n=124) and missing data on vital status (n=1). Reasons for not undergoing fundus photography (n = 969) included insufficient time (n = 539), physical limitation (n = 160), eye-specific limitation (n = 24), participant’s refusal (n = 110), communication problems (n = 26), and others (n=110). Excluded participants were more likely to be older, black, poorly educated, unmarried or other, have a lower level of household income, be unhealthier in terms of health-related behaviors and systemic comorbidities, and have a greater risk of mortality when compared to those with complete data. The characteristics of the excluded and the included participants are shown in Supplement Table 1.

A total of 710 participants were identified as having retinopathy (weighted prevalence: 9.75%). Among them, 573 (86.2%) out of 710 participants had minimal-to-mild NPR, 106 (11.2%) had moderate-to-severe NPR and 31 (2.65%) had PR. The mean age of these participants was 56.5 years (SE, 0.38), and 52.6% were women. The baseline characteristics of participants by retinopathy status are presented in Table 1. Participants identified as having retinopathy were more likely to be older, male, black, poorly educated, and lifetime abstainers/former drinkers and to have higher BMI, walking disability, poor/fair self-rated health status, and concomitant micro- and macrovascular pathologies (DM, HBP, CKD and CVD) compared to those without retinopathy. No significant difference in other characteristics was observed between these two groups. The baseline characteristics of participants stratified by retinopathy severity are shown in Supplement Table 2.

**Table 1.**
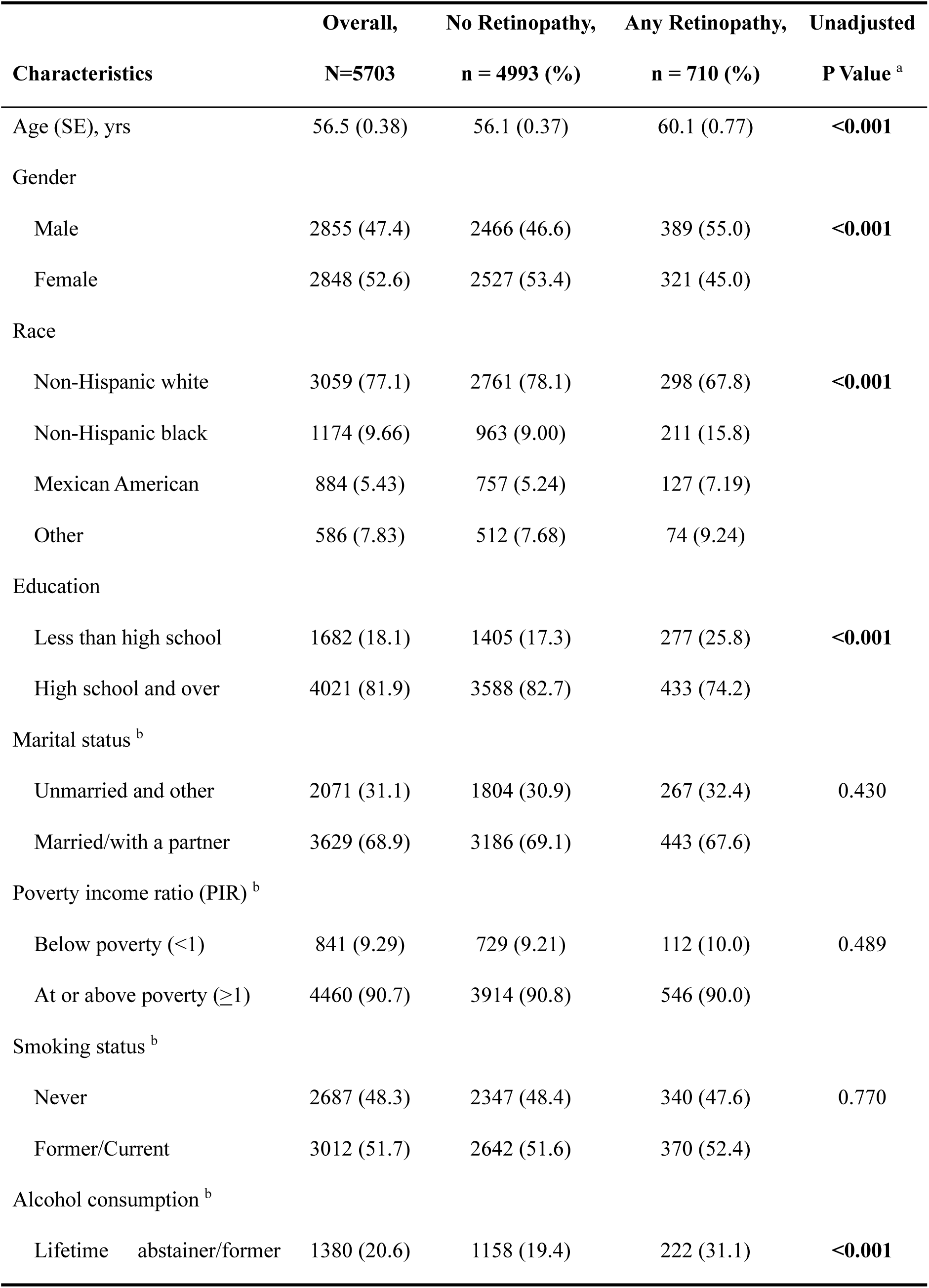
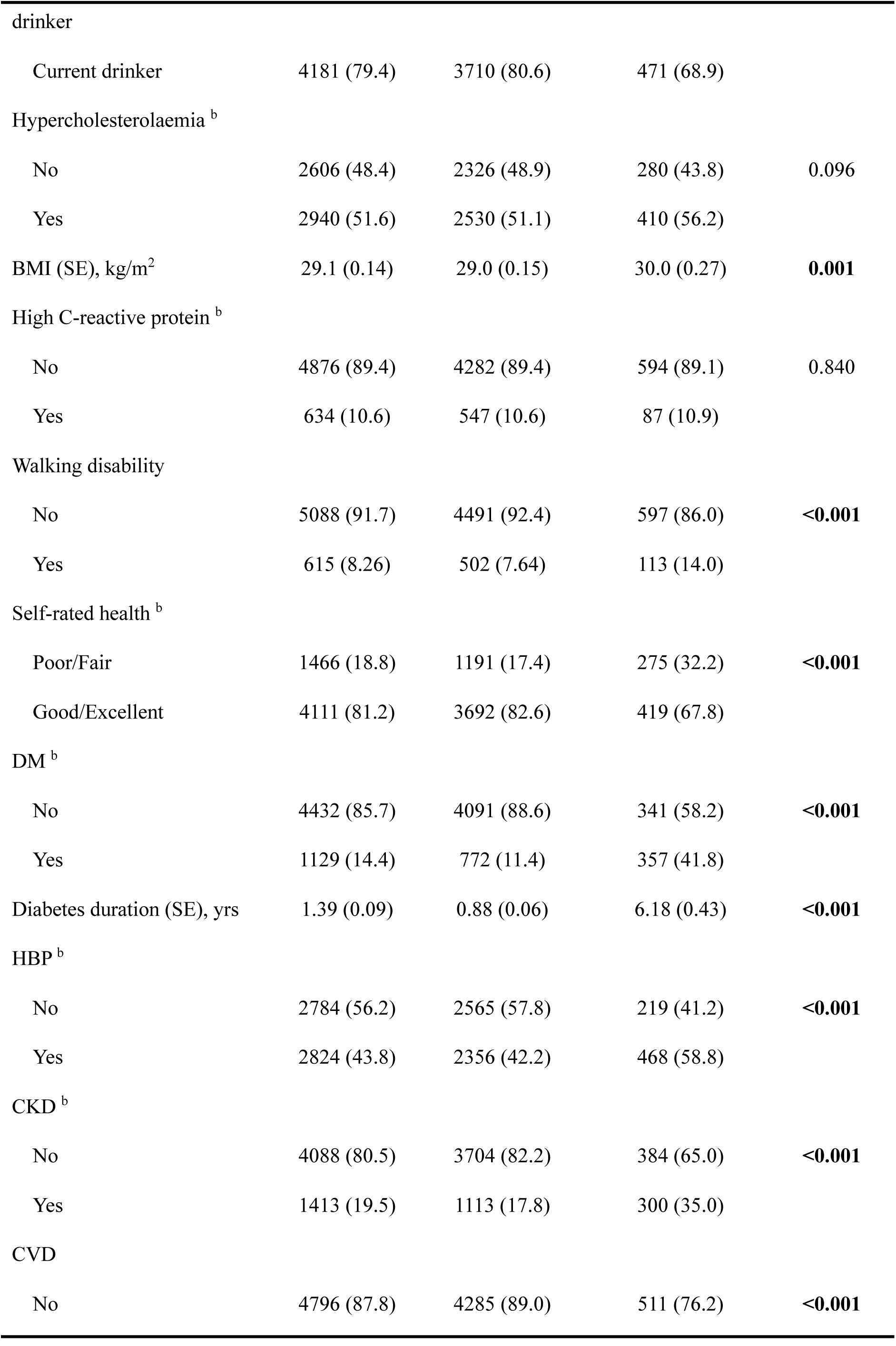
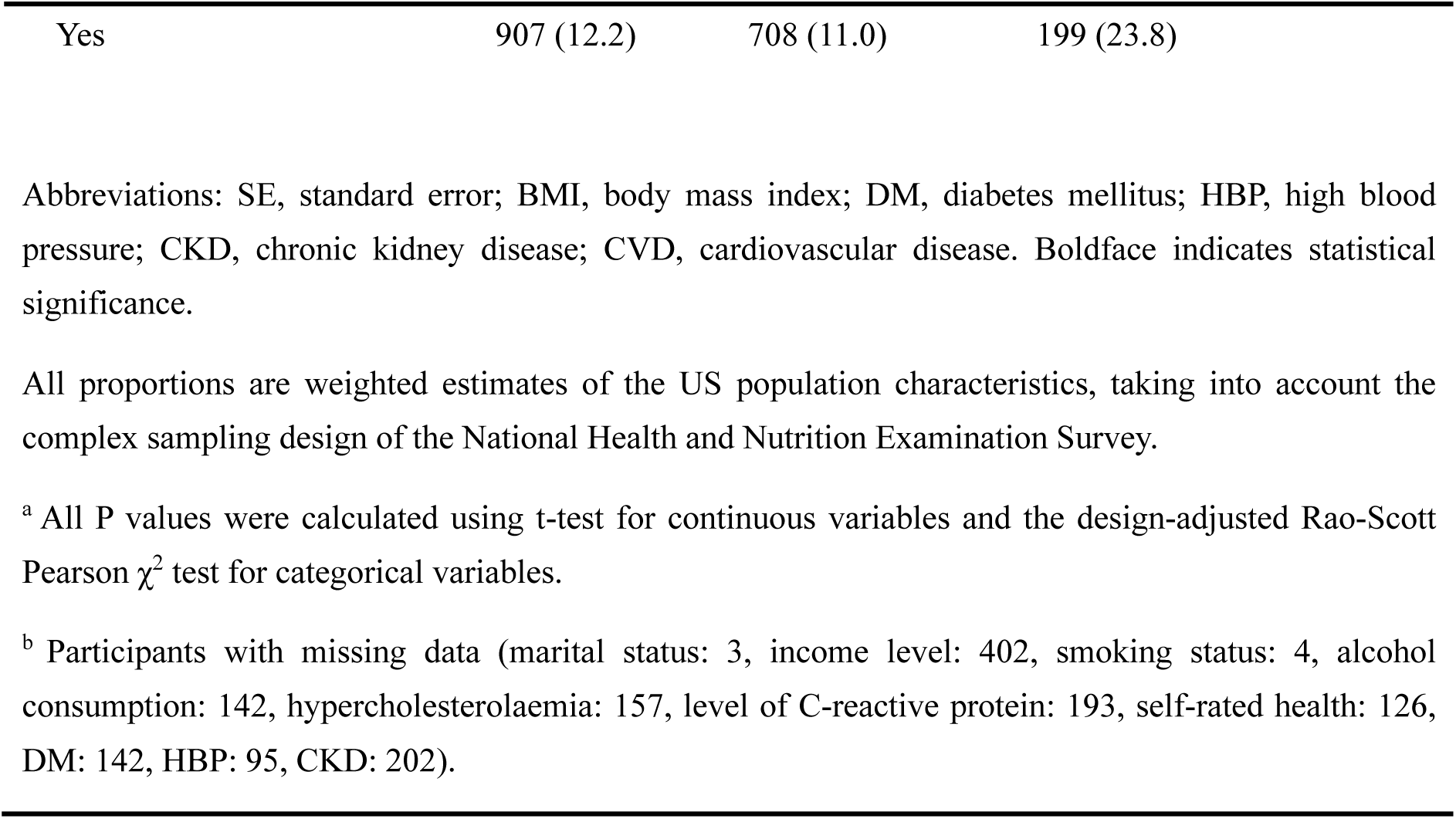
Demographic, Health-Related Behaviors and General Health Characteristics of Participants With and Without Retinopathy.

**Table 2.**
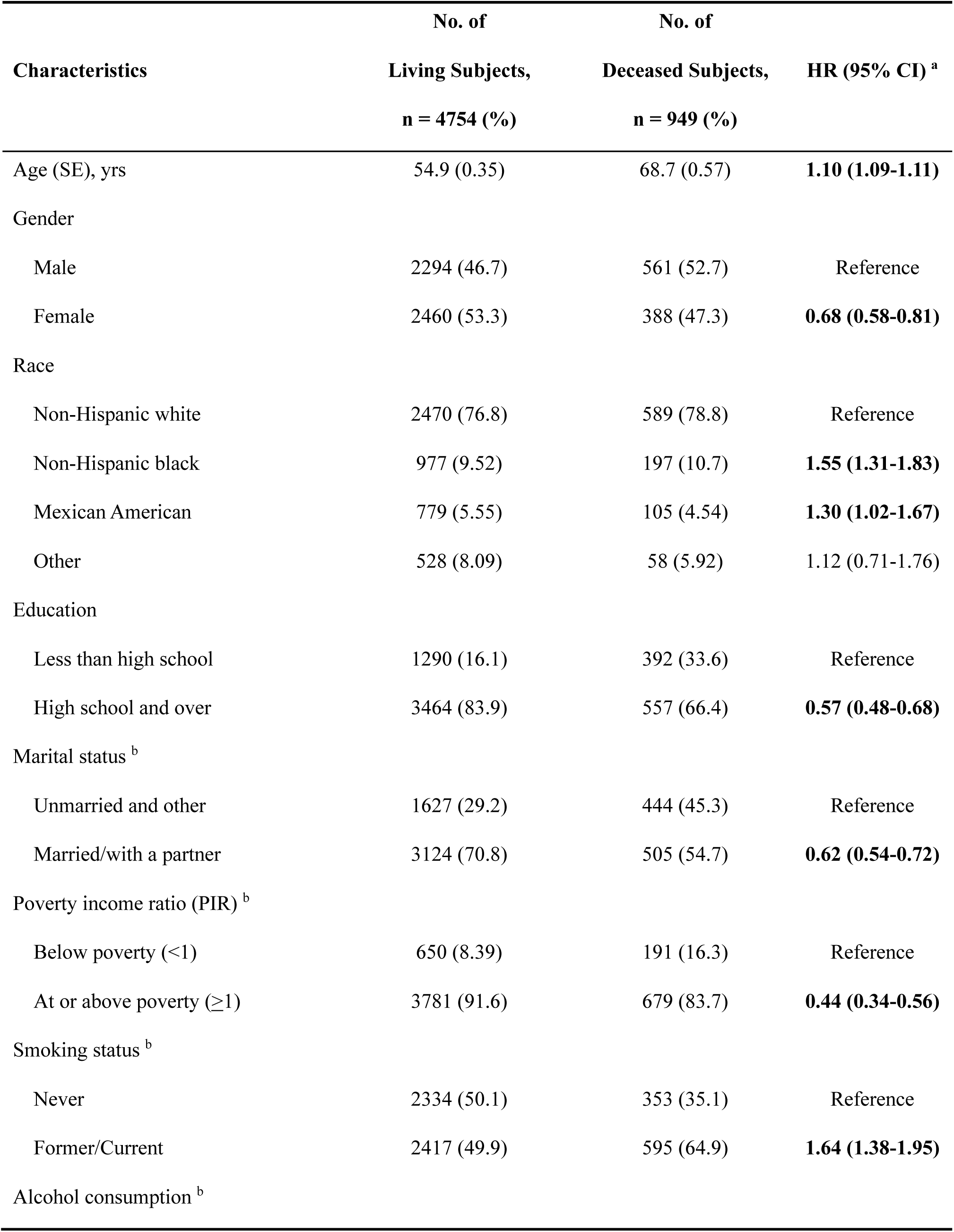
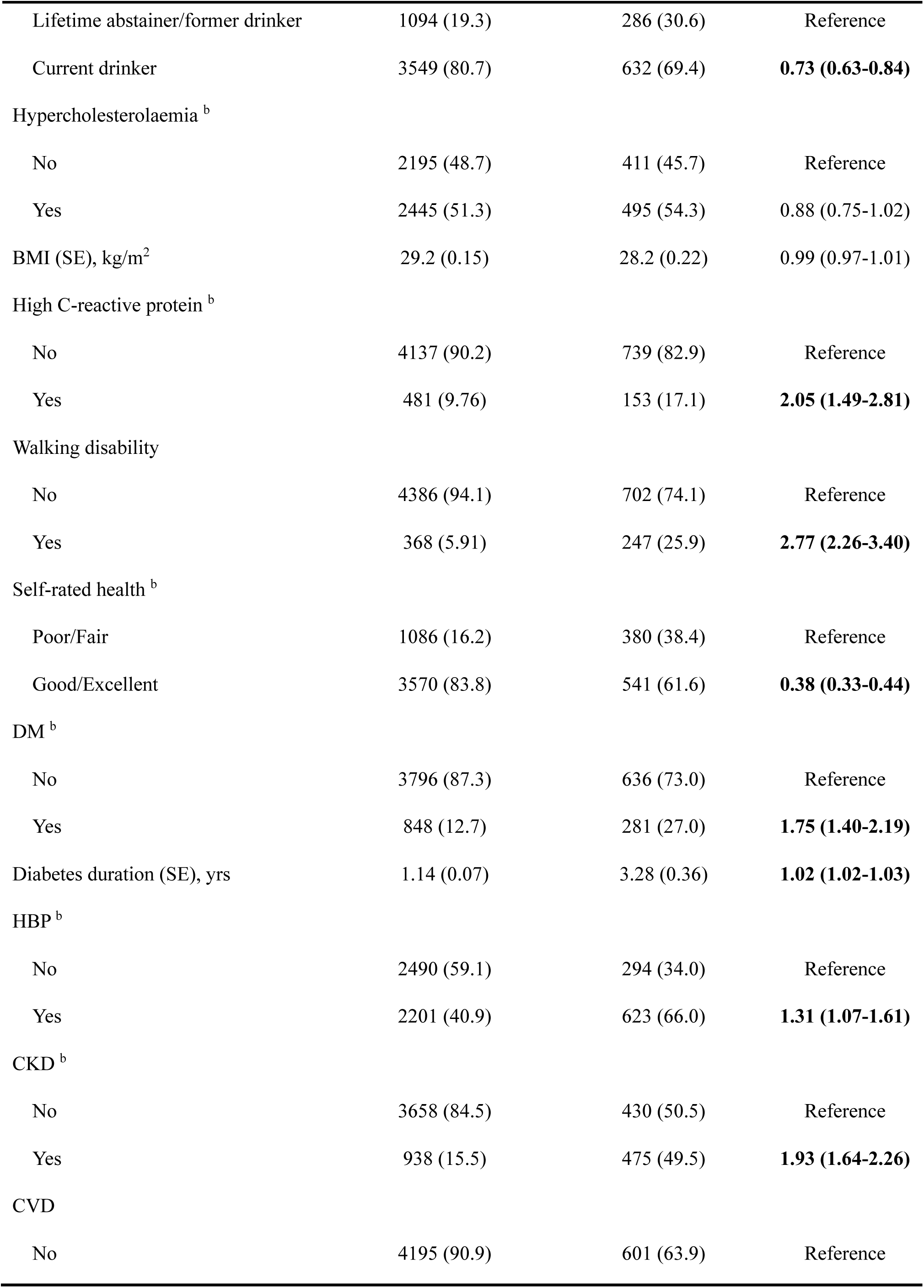
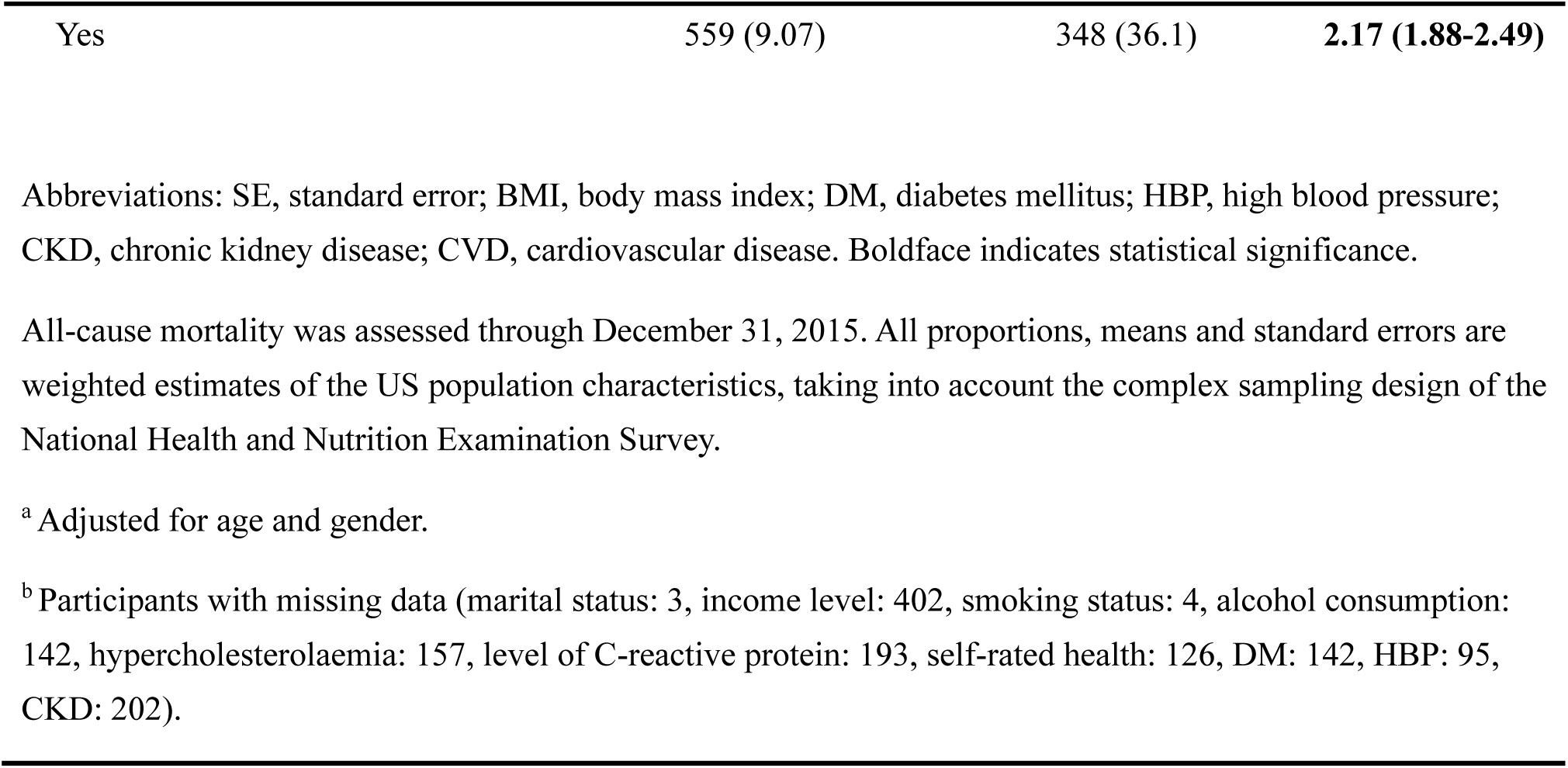
All-Cause Mortality by Demographic, Health-Related Behaviors and General Health Characteristics.

| Characteristics | No. of<br>Living Subjects,<br>n = 4754 (%) | No. of<br>Deceased Subjects,<br>n = 949 (%) | HR (95% CI) <sup>a</sup> |
| --- | --- | --- | --- |
| Age (SE), yrs | 54.9 (0.35) | 68.7 (0.57) | **1.10 (1.09-1.11)** |
| Gender |  |  |  |
| Male | 2294 (46.7) | 561 (52.7) | Reference |
| Female | 2460 (53.3) | 388 (47.3) | **0.68 (0.58-0.81)** |
| Race |  |  |  |
| Non-Hispanic white | 2470 (76.8) | 589 (78.8) | Reference |
| Non-Hispanic black | 977 (9.52) | 197 (10.7) | **1.55 (1.31-1.83)** |
| Mexican American | 779 (5.55) | 105 (4.54) | **1.30 (1.02-1.67)** |
| Other | 528 (8.09) | 58 (5.92) | 1.12 (0.71-1.76) |
| Education |  |  |  |
| Less than high school | 1290 (16.1) | 392 (33.6) | Reference |
| High school and over | 3464 (83.9) | 557 (66.4) | **0.57 (0.48-0.68)** |
| Marital status <sup>b</sup> |  |  |  |
| Unmarried and other | 1627 (29.2) | 444 (45.3) | Reference |
| Married/with a partner | 3124 (70.8) | 505 (54.7) | **0.62 (0.54-0.72)** |
| Poverty income ratio (PIR) <sup>b</sup> |  |  |  |
| Below poverty (<1) | 650 (8.39) | 191 (16.3) | Reference |
| At or above poverty ($\geq 1$) | 3781 (91.6) | 679 (83.7) | **0.44 (0.34-0.56)** |
| Smoking status <sup>b</sup> |  |  |  |
| Never | 2334 (50.1) | 353 (35.1) | Reference |
| Former/Current | 2417 (49.9) | 595 (64.9) | **1.64 (1.38-1.95)** |
| Alcohol consumption <sup>b</sup> |  |  |  |
| Lifetime abstainer/former drinker | 1094 (19.3) | 286 (30.6) | Reference |
| Current drinker | 3549 (80.7) | 632 (69.4) | **0.73 (0.63-0.84)** |
| Hypercholesterolaemia <sup>b</sup> |  |  |  |
| No | 2195 (48.7) | 411 (45.7) | Reference |
| Yes | 2445 (51.3) | 495 (54.3) | 0.88 (0.75-1.02) |
| BMI (SE), kg/m<sup>2</sup> | 29.2 (0.15) | 28.2 (0.22) | 0.99 (0.97-1.01) |
| High C-reactive protein <sup>b</sup> |  |  |  |
| No | 4137 (90.2) | 739 (82.9) | Reference |
| Yes | 481 (9.76) | 153 (17.1) | **2.05 (1.49-2.81)** |
| Walking disability |  |  |  |
| No | 4386 (94.1) | 702 (74.1) | Reference |
| Yes | 368 (5.91) | 247 (25.9) | **2.77 (2.26-3.40)** |
| Self-rated health <sup>b</sup> |  |  |  |
| Poor/Fair | 1086 (16.2) | 380 (38.4) | Reference |
| Good/Excellent | 3570 (83.8) | 541 (61.6) | **0.38 (0.33-0.44)** |
| DM <sup>b</sup> |  |  |  |
| No | 3796 (87.3) | 636 (73.0) | Reference |
| Yes | 848 (12.7) | 281 (27.0) | **1.75 (1.40-2.19)** |
| Diabetes duration (SE), yrs | 1.14 (0.07) | 3.28 (0.36) | **1.02 (1.02-1.03)** |
| HBP <sup>b</sup> |  |  |  |
| No | 2490 (59.1) | 294 (34.0) | Reference |
| Yes | 2201 (40.9) | 623 (66.0) | **1.31 (1.07-1.61)** |
| CKD <sup>b</sup> |  |  |  |
| No | 3658 (84.5) | 430 (50.5) | Reference |
| Yes | 938 (15.5) | 475 (49.5) | **1.93 (1.64-2.26)** |
| CVD |  |  |  |
| No | 4195 (90.9) | 601 (63.9) | Reference |
| Yes | 559 (9.07) | 348 (36.1) | **2.17 (1.88-2.49)** |

Abbreviations: SE, standard error; BMI, body mass index; DM, diabetes mellitus; HBP, high blood pressure; CKD, chronic kidney disease; CVD, cardiovascular disease. Boldface indicates statistical significance.
All-cause mortality was assessed through December 31, 2015. All proportions, means and standard errors are weighted estimates of the US population characteristics, taking into account the complex sampling design of the National Health and Nutrition Examination Survey.
<sup>a</sup> Adjusted for age and gender.
<sup>b</sup> Participants with missing data (marital status: 3, income level: 402, smoking status: 4, alcohol consumption: 142, hypercholesterolaemia: 157, level of C-reactive protein: 193, self-rated health: 126, DM: 142, HBP: 95, CKD: 202).

The median duration of follow-up was 8.33 years (interquartile range [IQR]: 7.50-9.67 years). There were 949 (11.8%) deaths from all causes. Participants with retinopathy had higher all-cause mortality than those without (23.0% versus 10.5%, t-test P < 0.001). The baseline characteristics, including demographic factors, health-related behaviors and general health characteristics for participants by survival status, are presented in Table 2. Several covariates, including age, sex, ethnicity, educational attainment, types of marital status, household income level, smoking status, alcohol consumption, the level of CRP, self-rated health status, walking disability, and micro- and macrovascular disorders (DM, CKD and history of CVD), were strongly associated with higher all-cause mortality in age- and sex-adjusted models. Multivariate Cox regression models suggested that the presence of any degree of retinopathy at baseline was associated with higher all-cause mortality (HR, 1.40; 95% CI, 1.09-1.81; P=0.011). There was a significant trend toward higher all-cause mortality risk across the retinopathy severity groups (P for trend = 0.010), where the greatest risk for all-cause mortality was among those with moderate-to-severe NPR or PR (HR=1.77, 95% CI: 1.04-3.03, P = 0.037).

Interactions between retinopathy and each systemic vascular disorder were assessed, and no significant interaction for DM, CKD, or CVD was found. There was a significant interaction between retinopathy and HBP in the model (P=0.045). Kaplan-Meier curves for all-cause mortality according to retinopathy and systemic vascular conditions are shown in Figure 1. Table 3 presents the synergistic impact of each vascular disorder together with retinopathy on mortality. All-cause mortality for participants with retinopathy and comorbidities such as DM (HR, 1.63; 95% CI, 1.06-2.50; P=0.027), HBP (HR, 1.46; 95% CI, 1.03-2.08; P=0.036), CKD (HR, 1.71; 95% CI, 1.24-2.35; P=0.002) and CVD (HR, 1.88; 95% CI, 1.19-2.96; P=0.008) was significantly higher than for those without any condition. The additional effects from vascular comorbidities on all-cause mortality are shown in Supplement Table 3. Overall, co-presence of retinopathy and vascular comorbidities further increased the risk of all-cause mortality than the presence of either health condition, but the increases in risks did not reach statistical significance.

**Table 3.**
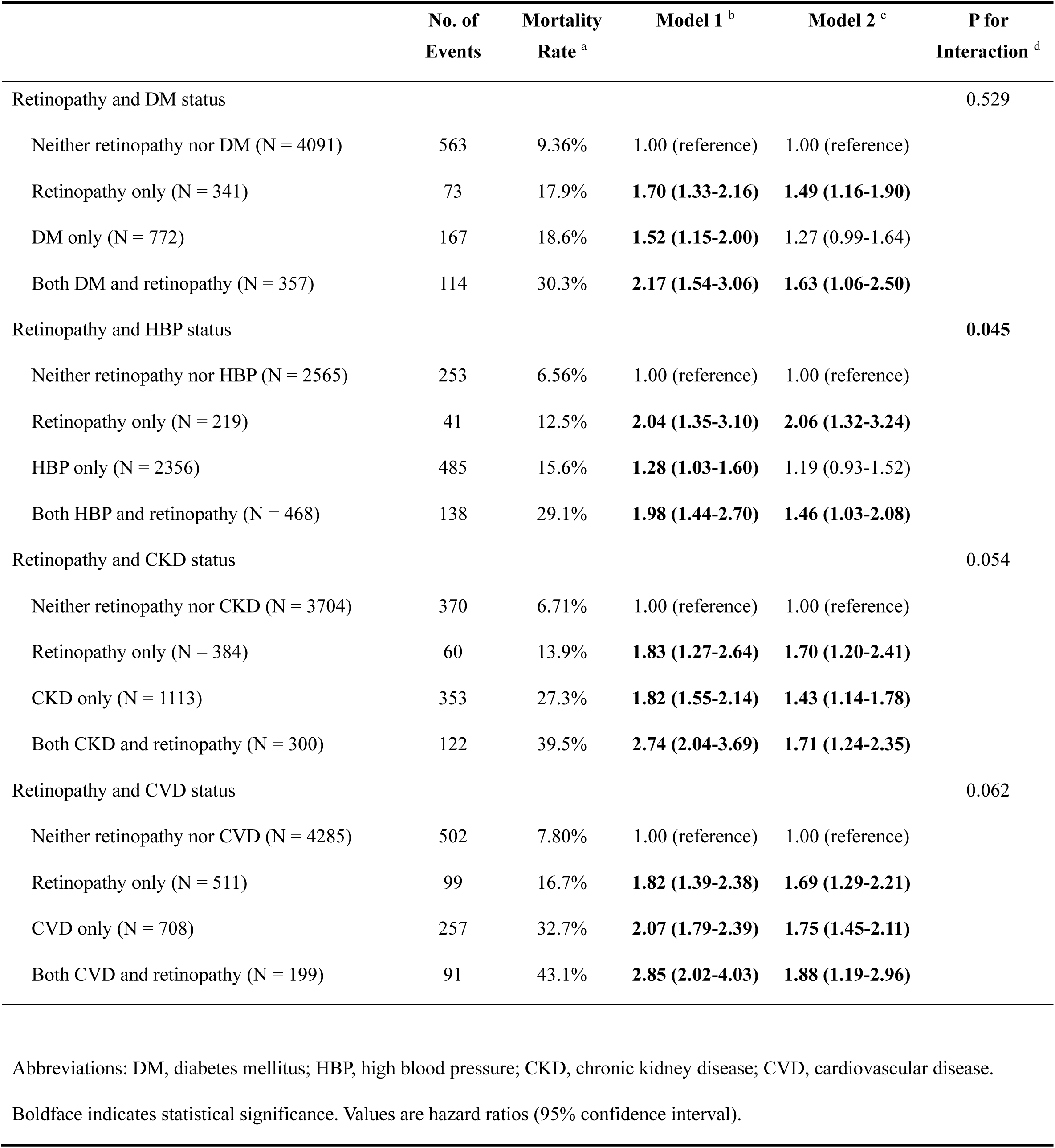
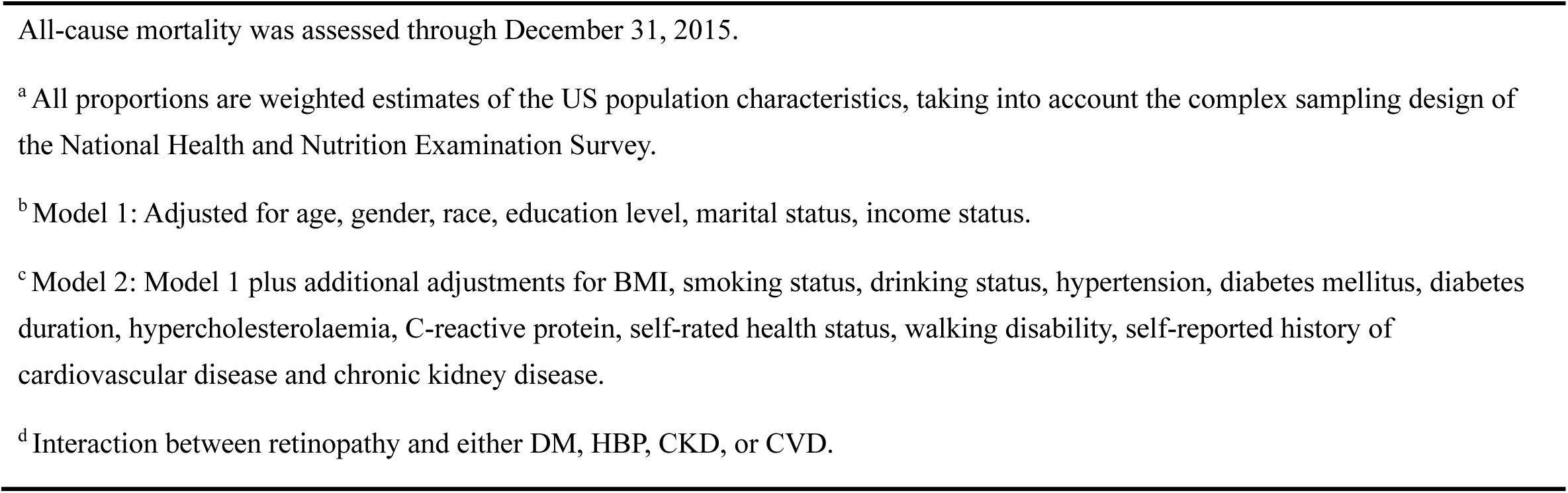
Cox Proportional Hazards Regression Models of All-Cause Mortality by Retinopathy Status and Concomitant Medical Conditions.

**Figure 1.**
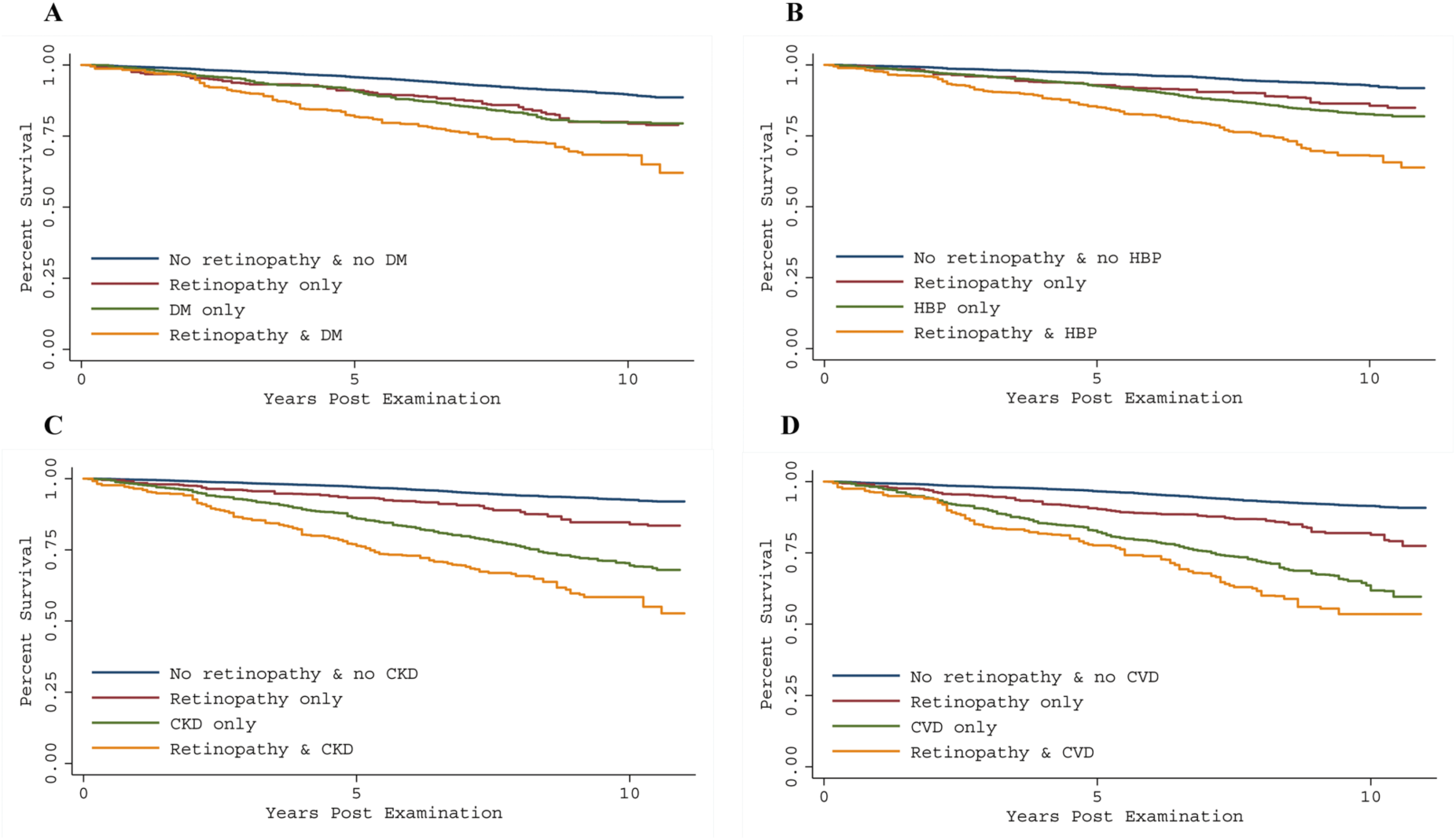
Kaplan-Meier curve showing all-cause mortality rate by retinopathy status and concomitant systemic vascular disorders (A: diabetes mellitus; B: high blood pressure; C: chronic kidney disease; D: history of cardiovascular disease), using the 2005-2008 National Health and Nutrition Examination Survey (NHANES). All-cause mortality was assessed through December 31, 2015. Risk of mortality was further increased among those with both retinopathy and systemic vascular diseases compared to participants without either condition.

The results stratified by diabetes and hypertension status are shown in Supplement Table 4. Among non-diabetic participants, all-cause mortality was higher in those with retinopathy and CKD (HR, 1.55; 95% CI, 1.07-2.25; P=0.023). Among diabetic participants, the co-occurrence of retinopathy and CVD (HR, 2.41; 95% CI, 1.15-5.08; P=0.022) further increased all-cause mortality. For participants without hypertension, the co-occurrence of retinopathy and CVD posed a higher risk of death (HR, 3.41; 95% CI, 1.35-8.61; P=0.011). In an analysis limited to participants with hypertension, the joint effect of retinopathy and CKD (HR, 1.83; 95% CI, 1.31-2.54; P=0.001) predicted higher all-cause mortality than that of those without either condition. The sensitivity analysis used inverse probability weighting for addressing the potential non-response bias, which led to similar results as to those reported in the main analysis (Supplement Table 5-7).

## Discussion

In this large sample of middle-aged and older adults, participants with retinopathy had a higher all-cause mortality rate. Moreover, the co-occurrence of retinopathy and systemic vascular conditions (DM, HBP, CKD, and history of CVD) further increased all-cause mortality.

Our findings are in line with previous results that reported a significant association between retinopathy and mortality.^17-26^ Contrary to the EURODIAB Prospective Complications Study, which found that the relationship between retinopathy and mortality can be largely explained by the presence of coexisting cardiovascular risk factors,^39^ we report that retinopathy was an independent risk factor for mortality after comprehensive adjustment of covariates. We postulate that the observed difference might be a result of missing fundus photographs (31%) and morbidity data (14%) in the EURODIAB Prospective Complications Study, which may have led to the underestimation of the association between retinopathy and mortality.

Only a limited number of studies have attempted to explore the joint effects of retinopathy and systemic vascular comorbidities (such as DM,^16^ CKD,^16, 29-31^ and clinical cerebrovascular disease^16^) on mortality. A more detailed investigation of systemic vascular comorbidities (DM, HBP, CKD, and CVD) in our study provides further insights into the association between retinopathy and mortality. That is, our results not only support previous findings of a higher mortality risk in individuals with retinopathy but also suggest that retinopathy in the presence of concomitant DM, HBP, CKD or CVD increases the risk of mortality even further.

Mechanisms underlying the association between retinopathy and mortality remain unknown. It has been proposed that retinopathy may be an indicator of the burden of CVD risk factors, including dyslipidemia, obesity, hypertension, and smoking.^17^ However, our results do not support this theory. Although we found that the distribution of CVD risk factors differed between the no retinopathy and retinopathy groups, retinopathy was found to be an independent risk factor for mortality in the fully adjusted model. Another potential explanation is that retinopathy, as a marker of abnormal microcirculation in the retina, may be an important indicator of systemic micro- and macro-vascular abnormalities.^24^ These micro- and macrovascular disorders may share similar pathophysiological processes with retinopathy, including endothelial dysfunction, inflammation, apoptosis, and neovascularization.^40, 41^ In addition, the co-occurrence of retinopathy and systemic vascular disorders may reflect a greater burden and/or severity of vascular pathologies, which may explain the further increase in mortality among the participants who have both retinopathy and systemic vascular disorders.

Given that retinopathy can be readily detected via non-invasive means, our findings may have several practical implications. Firstly, our finding of a statistically significant association between retinopathy and systemic vascular abnormalities indicates that individuals with retinopathy may benefit from a comprehensive vascular assessment and should be closely monitored. Secondly, the evidence of a further increase risk of mortality among patients with retinopathy and micro- or macrovascular disorders suggests that intensive vascular risk reduction is needed in the management of these patients. Last but not least, our results are of clinical and public health significance in the context of ageing population, highlighting the importance of retinopathy screening. Additionally, our evidence suggests that retinal imaging could be useful for identifying individuals at high risk of mortality, particularly among individuals with systemic vascular comorbidities, which could expand benefits of retinopathy screening beyond the prevention of sight-threatening disease. Of note, current guidelines and procedures vary across countries on who, when and how to conduct retinopathy screening.^42, 43^ The nationwide screening programs are limited.^44-46^ Nevertheless, the expanding role of retinopathy screening noted in the present analysis, together with the emerging of new technologies, such as tele-ophthalmology, portable fundus camera, and artificial intelligence (AI)-based automatic retinal image grading, are improving the cost-effectiveness of the nationwide screening programs.

The strengths of this analysis include the use of a large-scale sample with national representativeness, the use of a standardized objective grading protocol to assess retinopathy, access to death records, long-term follow-up and inclusion of a comprehensive range of covariates. However, several potential limitations should also be considered. Firstly, retinopathy status and potential confounders adjusted for in our analysis were assessed on a single occasion. During the follow-up period, the behaviors and/or retinopathy status of the patients may have changed, which may have a direct impact on the outcome. Secondly, discrepancies may exist between the self-reported data and clinically measured data. This may cause underreporting of milder cases and subsequently lead to a bias in the analysis. Thirdly, participants with missing data tended to have systematically higher mortality risk and possibly more severe retinopathy, which might bias our findings. Nevertheless, we built models for the probability of inclusion and using inverse probability weighting for addressing the potential non-response bias in the sensitivity analysis. Findings observed in the sensitivity analysis were consistent with those from the main analysis, thus verifying the robustness of our results. Fourthly, the number of deaths due to cardiovascular disease was relatively small in some subgroup (e.g., 10 in group with retinopathy but no CKD), prevented us from obtaining the robust estimation of joint effects of retinopathy and systemic vascular comorbidities on cardiovascular disease mortality. Further studies are needed to investigate joint effects of retinopathy and comorbidities on specific-cause mortality. Last but not least, we could not exclude the possibility of unmeasured or residual confounding by systemic comorbidities.

In summary, our findings suggest that middle-aged and elderly people with retinopathy have increased all-cause mortality. Furthermore, the joint effects of retinopathy and major systemic vascular comorbidities increase the all-cause mortality further. Our results indicate that more extensive risk factor assessment and management of individuals with retinopathy may be beneficial to reduce their mortality rate, especially in patients who have both retinopathy and systemic vascular disorders.

## Data Availability

The NHANES dataset used in this analysis were publicly available and de-identified, the present study received exemption from review by the institutional review board.

## Author Contributions

Study concept and design: Zhu ZT, He MG, Yang XH. Acquisition, analysis, or interpretation: All authors.

Drafting of the manuscript: Zhu ZT, Shang XW.

Critical revision of the manuscript for important intellectual content: Wang W, Jason H, Chen YF, He MG, Yang XH.

Statistical analysis: Zhu ZT, Shang XW, Wang W.

Obtained funding: He MG.

Administrative, technical, or material support: Zhu ZT, Shang XW, Wang W, He MG, Yang XH.

Study supervision: He MG, Yang XH.

## Abbreviations and Acronyms

CVD: cardiovascular disease
NHANES: National Health and Nutrition Examination Survey
DM: diabetes mellitus
HBP: high blood pressure
CKD: chronic kidney disease
HR: hazard ratios
CI: confidence interval
ETDRS: Early Treatment Diabetic Retinopathy Study
NDI: National Death Index
NPR: non-proliferative retinopathy
PR: proliferative retinopathy
eGFR: estimated glomerular filtration rate
ICD: International Classification of Diseases, Injuries and Causes of death
BMI: body mass index
CRP: C-reactive protein
PIR: poverty income ratio
SE: standard errors
VIF: variance inflation factors
IQR: interquartile range

Supplement Digital Content 1.

**Supplement Table 1**. Demographic, Health-Related Behaviors and General Health Characteristics of Participants Included and Excluded in the Analysis.

Supplement Digital Content 2.

**Supplement Table 2**. Stratified Analysis of Cox Proportional Hazards Regression Models of All-Cause Mortality by Retinopathy Status and Systemic Vascular Diseases.

Supplement Digital Content 3.

**Supplement Table 3**. Additional Effects from Vascular Comorbidities on All-Cause Mortality Using Cox Proportional Hazards Regression Models.

Supplement Digital Content 4.

**Supplement Table 4**. Stratified Analysis of Cox Proportional Hazards Regression Models of All-Cause Mortality by Retinopathy Status and Concomitant Medical Conditions.

Supplement Digital Content 5.

**Supplement Table 5**. Cox Proportional Hazards Regression Models of All-Cause Mortality by Retinopathy Status and Retinopathy Severity using Inverse Probability Weighting.

Supplement Digital Content 6.

**Supplement Table 6**. Cox Proportional Hazards Regression Models of All-Cause Mortality by Retinopathy Status and Concomitant Medical Conditions using Inverse Probability Weighting.

Supplement Digital Content 7.

**Supplement Table 7**. Stratified Analysis of Cox Proportional Hazards Regression Models of All-Cause by Retinopathy Status and Concomitant Medical Conditions using Inverse Probability Weighting.

