## Supplement Table 1 for "Impact of Retinopathy and Systemic Vascular Comorbidities on All-Cause Mortality"

Supplement Table 1. Demographic, Health-Related Behaviors and General Health Characteristics of Participants Included and Excluded in the Analysis

| **Characteristics** | **No. of Excluded Subjects, n = 1094 (%)** | **No. of Included Subjects, n = 5703 (%)** | **P Value [a]** |
|---|---|---|---|
| Age (SE), yrs | 60.9 (0.72) | 56.5 (0.38) | **<0.001** |
| Gender | | | |
| Male | 518 (45.0) | 2855 (47.4) | 0.226 |
| Female | 576 (55.0) | 2848 (52.6) | |
| Race | | | |
| Non-Hispanic white | 487 (65.7) | 3059 (77.1) | **<0.001** |
| Non-Hispanic black | 295 (16.1) | 1174 (9.66) | |
| Mexican American | 164 (6.16) | 884 (5.43) | |
| Other | 148 (12.0) | 586 (7.83) | |
| Education | | | |
| Less than high school | 459 (28.9) | 1682 (18.1) | **<0.001** |
| High school and over | 635 (71.1) | 4021 (81.9) | |
| Marital status | | | |
| Unmarried and other | 505 (38.8) | 2071 (31.1) | **0.001** |
| Married/with a partner | 586 (61.2) | 3629 (68.9) | |
| Poverty income ratio (PIR) | | | |
| Below poverty (<1) | 203 (13.9) | 841 (9.29) | **0.008** |
| At or above poverty (≥1) | 764 (86.1) | 4460 (90.7) | |
| Smoking status | | | |
| Never | 608 (55.6) | 2687 (48.3) | **0.002** |
| Former/Current | 483 (44.4) | 3012 (51.7) | |
| Alcohol consumption | | | |
| Lifetime abstainer/former drinker | 255 (34.1) | 1380 (20.6) | **<0.001** |

| | | | |
|---|---|---|---|
| Current drinker | 426 (65.9) | 4181 (79.4) | |
| Hypercholesterolaemia | | | |
| No | 456 (47.5) | 2606 (48.4) | 0.632 |
| Yes | 502 (52.5) | 2940 (51.6) | |
| BMI (SE), kg/m$^2$ | 28.5 (0.27) | 29.1 (0.14) | 0.117 |
| High C-reactive protein | | | |
| No | 796 (88.2) | 4876 (89.4) | 0.418 |
| Yes | 123 (11.8) | 634 (10.6) | |
| Walking disability | | | |
| No | 807 (78.0) | 5088 (91.7) | **<0.001** |
| Yes | 287 (22.0) | 615 (8.26) | |
| Self-rated health | | | |
| Poor/Fair | 264 (31.2) | 1466 (18.8) | **<0.001** |
| Good/Excellent | 428 (68.8) | 4111 (81.2) | |
| DM | | | |
| No | 669 (75.1) | 4432 (85.7) | **<0.001** |
| Yes | 295 (24.9) | 1129 (14.4) | |
| Diabetes duration (SE), yrs | 3.30 (0.34) | 1.39 (0.09) | **<0.001** |
| HBP | | | |
| No | 349 (46.1) | 2784 (56.2) | **<0.001** |
| Yes | 515 (53.9) | 2824 (43.8) | |
| CKD | | | |
| No | 507 (64.3) | 4088 (80.5) | **<0.001** |
| Yes | 385 (35.7) | 1413 (19.5) | |
| CVD | | | |
| No | 833 (79.6) | 4796 (87.8) | **<0.001** |
| Yes | 261 (20.4) | 907 (12.2) | |

| | | | |
|---|---|---|---|
| Vital status | | | |
| Survived | 715 (71.1) | 4754 (88.3) | **<0.001** |
| Deceased | 377 (28.9) | 949 (11.7) | |

Abbreviations: SE, standard error; BMI, body mass index; DM, diabetes mellitus; HBP, high blood pressure; CKD, chronic kidney disease; CVD, cardiovascular disease.

All proportions are weighted estimates of the US population characteristics, taking into account the complex sampling design of the National Health and Nutrition Examination Survey.

[a] All P values were calculated using the t-test for continuous variables and design-adjusted Rao-Scott Pearson $\chi^2$ test for categorical variables. Boldface indicates statistical significance.
