## Supplement Table 2 for "Impact of Retinopathy and Systemic Vascular Comorbidities on All-Cause Mortality"

Supplement Table 2. Demographic, Health-Related Behaviors and General Health Characteristics of Participants Stratified by Retinopathy Severity.

| **Characteristics** | **No Retinopathy, n=4993 (%)** | **Minimal-to-Mild NPR, n=573 (%)** | **Moderate-to-Severe NPR/PR, n=137 (%)** | **Unadjusted P Value** [a] |
|---|---|---|---|---|
| Age (SE), yrs | 56.1 (0.37) | 60.0 (0.85) | 60.4 (1.03) | **<0.001** |
| Gender | | | | |
| Male | 2466 (46.6) | 324 (55.7) | 65 (50.6) | **<0.001** |
| Female | 2527 (53.4) | 249 (44.3) | 72 (49.4) | |
| Race | | | | |
| Non-Hispanic white | 2761 (78.1) | 266 (70.4) | 32 (51.6) | **<0.001** |
| Non-Hispanic black | 963 (9.00) | 150 (13.3) | 61 (31.2) | |
| Mexican American | 757 (5.24) | 94 (6.34) | 33 (12.5) | |
| Other | 512 (7.68) | 63 (9.96) | 11 (4.76) | |
| Education | | | | |
| Less than high school | 1405 (17.3) | 214 (24.5) | 63 (34.1) | **<0.001** |
| High school and over | 3588 (82.7) | 359 (75.5) | 74 (65.9) | |
| Marital status | | | | |
| Unmarried and other | 1804 (30.9) | 208 (31.6) | 59 (37.5) | 0.370 |
| Married/with a partner | 3186 (69.1) | 365 (68.4) | 78 (62.5) | |
| Poverty income ratio (PIR) | | | | |
| Below poverty (<1) | 729 (9.21) | 83 (8.80) | 29 (18.0) | **0.049** |
| At or above poverty (≥1) | 3914 (90.8) | 453 (91.2) | 93 (82.0) | |
| Smoking status | | | | |
| Never | 2347 (48.4) | 264 (45.4) | 76 (61.9) | **0.045** |
| Former/Current | 2642 (51.6) | 309 (54.6) | 61 (38.1) | |
| Alcohol consumption | | | | |
| Lifetime abstainer/former drinker | 1158 (19.4) | 172 (30.7) | 50 (33.7) | **<0.001** |

| | | | | |
|---|---|---|---|---|
| Current drinker | 3710 (80.6) | 386 (69.3) | 85 (66.3) | |
| Hypercholesterolaemia | | | | |
| No | 2326 (48.9) | 237 (45.9) | 43 (30.5) | **0.021** |
| Yes | 2530 (51.1) | 325 (54.1) | 85 (69.5) | |
| BMI (SE), kg/m$^2$ | 29.0 (0.15) | 29.4 (0.29) | 33.4 (1.01) | **<0.001** |
| High C-reactive protein | | | | |
| No | 4282 (89.4) | 495 (91.0) | 99 (77.1) | **0.008** |
| Yes | 547 (10.6) | 60 (9.02) | 27 (22.9) | |
| Walking disability | | | | |
| No | 4491 (92.4) | 495 (87.6) | 102 (76.0) | **<0.001** |
| Yes | 502 (7.64) | 78 (12.4) | 35 (24.0) | |
| Self-rated health | | | | |
| Poor/Fair | 1191 (17.4) | 199 (29.3) | 76 (49.9) | **<0.001** |
| Good/Excellent | 3692 (82.6) | 360 (70.7) | 59 (50.1) | |
| DM | | | | |
| No | 4091 (88.6) | 331 (66.5) | 10 (7.55) | **<0.001** |
| Yes | 772 (11.4) | 231 (33.5) | 126 (92.5) | |
| Diabetes duration (SE), yrs | 0.88 (0.06) | 4.59 (0.40) | 16.0 (1.21) | **<0.001** |
| HBP | | | | |
| No | 2565 (57.8) | 190 (43.3) | 29 (27.8) | **<0.001** |
| Yes | 2356 (42.2) | 366 (56.7) | 102 (72.2) | |
| CKD | | | | |
| No | 3704 (82.2) | 338 (69.0) | 46 (38.8) | **<0.001** |
| Yes | 1113 (17.8) | 218 (31.0) | 82 (61.2) | |
| CVD | | | | |
| No | 4285 (89.0) | 429 (77.7) | 82 (66.6) | **<0.001** |
| Yes | 708 (11.0) | 144 (22.3) | 55 (33.4) | |

Abbreviations: SE, standard error; BMI, body mass index; DM, diabetes mellitus; HBP, high blood pressure; CKD, chronic kidney disease; CVD, cardiovascular disease. Boldface indicates statistical significance.

All proportions are weighted estimates of the US population characteristics, taking into account the complex sampling design of the National Health and Nutrition Examination Survey.

[a] All P values were calculated using t-test for continuous variables and the design-adjusted Rao-Scott Pearson $\chi^2$ test for categorical variables.
