## Supplement Table 3 for "Impact of Retinopathy and Systemic Vascular Comorbidities on All-Cause Mortality"

Supplement Table 3. Additional Effects from Vascular Comorbidities on All-Cause Mortality Using Cox Proportional Hazards Regression Models.

| | **Model 1** [a] | **Model 2** [b] |
|---|---|---|
| Retinopathy and DM status | | |
| Both DM and retinopathy v.s. Retinopathy only | 1.28 (0.86-1.88) | 1.10 (0.68-1.76) |
| Both DM and retinopathy v.s. DM only | 1.43 (0.92-2.21) | 1.28 (0.82-2.01) |
| Retinopathy and HBP status | | |
| Both HBP and retinopathy v.s. Retinopathy only | 0.97 (0.63-1.49) | 0.71 (0.44-1.14) |
| Both HBP and retinopathy v.s. HBP only | **1.54 (1.17-2.03)** | 1.23 (0.92-1.63) |
| Retinopathy and CKD status | | |
| Both CKD and retinopathy v.s. Retinopathy only | 1.50 (0.99-2.25) | 1.00 (0.66-1.54) |
| Both CKD and retinopathy v.s. CKD only | **1.51 (1.15-1.96)** | 1.20 (0.92-1.56) |
| Retinopathy and CVD status | | |
| Both CVD and retinopathy v.s. Retinopathy only | **1.57 (1.05-2.35)** | 1.11 (0.69-1.78) |
| Both CVD and retinopathy v.s. CVD only | 1.38 (0.96-1.99) | 1.07 (0.70-1.64) |

Abbreviations: DM, diabetes mellitus; HBP, high blood pressure; CKD, chronic kidney disease; CVD, cardiovascular disease.

Boldface indicates statistical significance. Values are hazard ratios (95% confidence interval).

All-cause mortality was assessed through December 31, 2015.

[a] Model 1: Adjusted for age, gender, race, education level, marital status, income status.

[b] Model 2: Model 1 plus additional adjustments for BMI, smoking status, drinking status, hypertension, diabetes mellitus, diabetes duration, hypercholesterolaemia, C-reactive protein, self-rated health status, walking disability, self-reported history of cardiovascular disease and chronic kidney disease.
