## Supplement Table 4 for "Impact of Retinopathy and Systemic Vascular Comorbidities on All-Cause Mortality"

Supplement Table 4. Stratified Analysis of Cox Proportional Hazards Regression Models of All-Cause Mortality by Retinopathy Status and Concomitant Medical Conditions.

| | **All-Cause Mortality** | | | |
|---|---|---|---|---|
| | **DM** | | **HBP** | |
| | **No (N = 4432)** | **Yes (N = 1129)** | **No (N = 2784)** | **Yes (N = 2824)** |
| Retinopathy and CKD status | | | | |
| Neither retinopathy nor CKD | 1.00 (reference) | 1.00 (reference) | 1.00 (reference) | 1.00 (reference) |
| Retinopathy only | **1.99 (1.32-2.99)** | 1.20 (0.51-2.80) | **1.85 (1.20-2.86)** | **1.64 (1.03-2.62)** |
| CKD only | **1.41 (1.12-1.76)** | 1.35 (0.80-2.29) | 0.81 (0.60-1.11) | **1.78 (1.32-2.39)** |
| Both CKD and retinopathy | **1.55 (1.07-2.25)** | 1.70 (0.90-3.20) | 2.10 (0.74-5.96) | **1.83 (1.31-2.54)** |
| Retinopathy and CVD status | | | | |
| Neither retinopathy nor CVD | 1.00 (reference) | 1.00 (reference) | 1.00 (reference) | 1.00 (reference) |
| Retinopathy only | **1.83 (1.31-2.56)** | 1.62 (0.77-3.41) | **2.07 (1.20-3.57)** | 1.40 (0.97-2.03) |
| CVD only | **1.51 (1.20-1.89)** | **2.50 (1.51-4.15)** | **1.94 (1.20-3.15)** | **1.59 (1.16-2.18)** |
| Both CVD and retinopathy | 1.56 (0.88-2.78) | **2.41 (1.15-5.08)** | **3.41 (1.35-8.61)** | 1.61 (0.97-2.69) |

Abbreviations: DM, diabetes millitus; HBP, high blood pressure; CKD, chronic kidney disease; CVD, cardiovascular disease.

Boldface indicates statistical significance. Values are number of hazard ratio (95% confidence interval).

All-cause mortality was assessed through December 31, 2015.

All proportions are weighted estimates of the US population characteristics, taking into account the complex sampling design of the National Health and Nutrition Examination Survey.

[a] Adjusted for age, gender, race, education level, marital status, income status, BMI, smoking status, drinking status, hypertension, diabetes millitus, diabetes duration, cholesterol level, C-reactive protein, self-rated health status, walking disability, self-reported history of cardiovascular disease and chronic kidney disease.
