## Supplement Table 5 for "Impact of Retinopathy and Systemic Vascular Comorbidities on All-Cause Mortality"

Supplement Table 5. Cox Proportional Hazards Regression Models of All-Cause Mortality by Retinopathy Status and Retinopathy Severity using Inverse Probability Weighting.

| | **Model 1 [a]** | **Model 2 [b]** |
|---|---|---|
| Retinopathy status | | |
| No retinopathy | 1.00 (reference) | 1.00 (reference) |
| Retinopathy | **1.77 (1.39-2.25)** | **1.37 (1.07-1.75)** |
| Retinopathy severity | | |
| No retinopathy | 1.00 (reference) | 1.00 (reference) |
| Minimal-to-mild NPR | **1.65 (1.27-2.14)** | **1.33 (1.04-1.71)** |
| Moderate-to-severe NPR or PR | **2.40 (1.60-3.58)** | **1.67 (1.02-2.71)** |

Abbreviations: NPR, non-proliferative retinopathy; PR, proliferative retinopathy.

Boldface indicates statistical significance. Values are hazard ratios (95% confidence interval).

All-cause mortality was assessed through December 31, 2015.

[a] Model 1: Adjusted for age, gender, race, education level, marital status, income status.

[b] Model 2: Model 1 plus additional adjustments for BMI, smoking status, drinking status, hypertension, diabetes mellitus, diabetes duration, hypercholesterolaemia, C-reactive protein, self-rated health status, walking disability, self-reported history of cardiovascular disease and chronic kidney disease.
