## Supplement Table 6 for "Impact of Retinopathy and Systemic Vascular Comorbidities on All-Cause Mortality"

Supplement Table 6. Cox Proportional Hazards Regression Models of All-Cause Mortality by Retinopathy Status and Concomitant Medical Conditions using Inverse Probability Weighting.

| | **Model 1** [a] | **Model 2** [b] |
|---|---|---|
| Retinopathy and DM status | | |
| Neither retinopathy nor DM | 1.00 (reference) | 1.00 (reference) |
| Retinopathy only | **1.66 (1.32-2.09)** | **1.47 (1.15-1.86)** |
| DM only | **1.53 (1.17-2.01)** | 1.27 (0.98-1.64) |
| Both DM and retinopathy | **2.21 (1.59-3.07)** | **1.58 (1.03-2.43)** |
| Retinopathy and HBP status | | |
| Neither retinopathy nor HBP | 1.00 (reference) | 1.00 (reference) |
| Retinopathy only | **2.22 (1.44-3.41)** | **2.08 (1.33-3.25)** |
| HBP only | **1.27 (1.01-1.59)** | 1.21 (0.95-1.54) |
| Both HBP and retinopathy | **2.01 (1.49-2.71)** | **1.45 (1.03-2.04)** |
| Retinopathy and CKD status | | |
| Neither retinopathy nor CKD | 1.00 (reference) | 1.00 (reference) |
| Retinopathy only | **1.86 (1.29-2.69)** | **1.68 (1.18-2.37)** |
| CKD only | **1.66 (1.40-1.96)** | **1.42 (1.14-1.76)** |
| Both CKD and retinopathy | **2.49 (1.86-3.33)** | **1.68 (1.24-2.27)** |
| Retinopathy and CVD status | | |
| Neither retinopathy nor CVD | 1.00 (reference) | 1.00 (reference) |
| Retinopathy only | **1.83 (1.40-2.39)** | **1.65 (1.27-2.14)** |
| CVD only | **2.07 (1.74-2.47)** | **1.76 (1.46-2.12)** |
| Both CVD and retinopathy | **2.88 (2.04-4.05)** | **1.87 (1.20-2.92)** |

Abbreviations: DM, diabetes mellitus; HBP, high blood pressure; CKD, chronic kidney disease; CVD, cardiovascular disease.

Boldface indicates statistical significance. Values are hazard ratios (95% confidence interval).

All-cause mortality was assessed through December 31, 2015.

[a] Model 1: Adjusted for age, gender, race, education level, marital status, income status.

[b] Model 2: Model 1 plus additional adjustments for BMI, smoking status, drinking status, hypertension, diabetes mellitus, diabetes duration, hypercholesterolaemia, C-reactive protein, self-rated health status, walking disability, self-reported history of cardiovascular disease and chronic kidney disease.
